# Serious psychological distress among people with unstable housing in Ho Chi Minh City, Vietnam: Prevalence and associated factors

**DOI:** 10.1101/2024.12.04.24318512

**Authors:** Hitoshi Murakami, Nguyen Thuy Linh, Masami Fujita, Lam Ngoc Thuy, Nguyen Hong Phuc, Kieu Thi Mai Huong, Le Tuan Anh, Pham Thi Ngoc Mai, Khuat Thi Hai Oanh

## Abstract

**Purpose:** This study aimed to estimate the prevalence of serious psychological distress (SPD) among slum dwellers and homeless individuals in Ho Chi Minh City, Vietnam, and to identify factors associated with SPD.

**Method:** A cross-sectional survey of 415 individuals with unstable housing, including 383 slum dwellers and 32 homeless individuals, was conducted from November 2023 to April 2024. Data were collected using a structured questionnaire that included the Kessler Psychological Distress Scale (K6).

**Results:** The prevalence of SPD was 19.8% (18.5% among slum dwellers and 34.4% among the homeless). Multiple logistic regression revealed significant associations between SPD and female gender (p=0.000), labor exploitation (p=0.046), and debt (p=0.000). Notably, 68.7% of participants reported experiencing some form of labor exploitation, commonly involving contract rejections, forced overwork, wage theft, and physical or verbal abuse. Additionally, 43.7% were in debt, with 38.6% borrowing from moneylenders.

**Conclusions:** The prevalence of SPD among individuals with unstable housing in Ho Chi Minh City was significantly higher than that of the general population. Those with SPD frequently faced both labor and economic exploitation, without insufficient social protection. In terms of labor exploitation, policy interventions, particularly from an occupational health perspective, are necessary. To address economic exploitation through debt, given the prevalence of loan sharks, efforts to crack down on predatory lending and promote financial inclusion are essential.

## Introduction

Housing is one of the major social determinants of health [1–4]. Unstable housing, whether among migrants or domestic residents, is known to be associated with mental health issues, including psychological distress, depression, anxiety, and suicide [5–10]. The relationship between unstable housing and mental health problems is reported to be mediated by various factors, including stressful life events, substance abuse [11], and childhood learning disabilities [12]. Poverty manifests itself on multiple levels, ranging from income inequality at the macro level to precarious housing, food insecurity, and malnutrition at the micro level [13, 14]. In fact, housing insecurity is not an isolated phenomenon but is interlinked with other deprivations stemming from poverty, such as food insecurity, energy poverty, and unemployment [15].

Ho Chi Minh City (HCMC) is the largest city in Vietnam, with a population of 9,456,700 as of 2023 [16], and serves as the country’s industrial center. Unstable housing has been recognized as a significant issue in HCMC for decades, but its exact scale has seldom been accurately measured [17]. In 2004, approximately 15% of the city’s housing consisted of slums or squatter settlements. These informal areas are primarily inhabited by low-income households, many of whom live in precarious conditions without adequate sanitation or housing stability. The city’s rapid urbanization, with thousands of new migrants arriving each year, has strained housing resources and contributed to the expansion of these settlements [18[. In 2002, municipal authorities had registered 67,000 households living in what residents call “rat holes,” referring to the precarious houses in slums, over one-third of which were built along the city’s canals [19]. In 1995, it was estimated that 5% of the city’s population was homeless [20, 21]. Given the city’s population in 1995 was 4,640,000 [22], this suggests that approximately 232,000 people were homeless at that time. In 2003-2004, more than 8,000 street children were living in HCMC [23]. However, as of 2019, only 39 homeless families were officially recorded in HCMC [24], a figure that was significantly underestimated as it excluded all individuals sleeping in roofed structures such as staircases, and did not account for housing ownership.

Systematic reviews have identified mental health as a significant issue among slum dwellers, particularly among women [25] and older individuals in Sub-Saharan Africa [26]. However, there is limited literature on mental health in slum settings or how it is influenced by the social environment in these neighborhoods [27]. These and other reviews highlight various social determinants of health related to slum living, such as socioeconomic status, gender, living conditions, food insecurity, social capital, social support [25], healthcare access and utilization [26], housing, nutrition, neighborhood characteristics, occupational factors, and health behaviors [28].

Reflecting the fact that homelessness is a significant social issue in high-income countries, whereas slum dwelling is primarily a problem in low- and middle-income countries (LMICs), pooled estimates of the prevalence of mental disorders among homeless populations have been derived from systematic reviews and meta-analyses [29]. The pooled prevalence of mental disorders among homeless individuals was estimated to be 9% in the United States, 12% in Western Europe, 19% in the United Kingdom, and 16% in Australia [30]. Another review, focusing on high-income countries, estimated that the mean prevalence of any current mental disorder was 76.2%, with the most common diagnostic categories being alcohol use disorders (36.7%) and drug use disorders (21.7%), followed by schizophrenia spectrum disorders (12.4%) and major depression (12.6%) [31]. A systematic review of psychopathology among homeless youth revealed that the prevalence of DSM and ICD disorders ranged from 48.4% to 98% [32]. In Germany, the pooled prevalence of mental illness among the homeless population was 77.5%, with substance-related disorders (60.9%)—particularly alcoholism (55.4%)—being the most common [33]. Among homeless school-age children in the United States, 24% to 40% had mental health problems, a rate two to four times higher than that of housed children living in poverty [34].

Psychological distress (PD) is generally defined as a state of emotional suffering, characterized by symptoms of depression (e.g., loss of interest, sadness, hopelessness) and anxiety (e.g., restlessness, tension) [35]. Rider provided a working definition of PD as the unique, discomforting emotional state experienced by an individual in response to a specific stressor or demand, which results in harm—either temporary or permanent—to the person [36]. Although PD is often described as a non-specific mental health issue [35, 37], as reflected in the definition above, it is clearly characterized by symptoms of depression and anxiety. Thus, while PD and these mental disorders are distinct phenomena, they are not entirely separate [38]. Scales measuring PD, such as the Kessler Psychological Distress Scales (K6/K10), have been reported to most effectively detect major depression and dysthymia according to the DSM-IV. Studies have shown that the K6/K10 can estimate the population prevalence of 12-month DSM-IV mood, anxiety, or substance use disorders with good accuracy in Australia [39]. The screening performance of the K6/K10 in detecting 30-day DSM-IV mood and anxiety disorders was also found to be excellent in Japan [40].

Building on the above background, the present study aimed to: 1) examine socio-economic vulnerabilities, 2) estimate the prevalence of serious psychological distress (SPD), and 3) identify social determinants of health significantly associated with SPD among populations with unstable housing, including slum dwellers and homeless individuals, in HCMC, Vietnam. The study was conducted by the Center for Supporting Community Development Initiatives (SCDI), Vietnam, in collaboration with the National Center for Global Health and Medicine, Japan. SCDI is a non-profit organization established in 2010, with the mission of improving the quality of life and promoting social inclusion for vulnerable and marginalized populations in Vietnam. SCDI has been actively supporting those with unstable housing, including slum dwellers and the homeless, in HCMC.

## Methods

### Target population

The target population of the present study comprised individuals with unstable housing, including those living in slums or experiencing homelessness, who were accessed by SCDI’s outreach team or sought its drop-in services in HCMC. The inclusion criteria were: 1) individuals present during household visits, 2) aged 18 years or older, and 3) those who could represent the household if more than one person was present. The exclusion criteria were: 1) individuals who did not agree to participate in the survey and 2) those who could not understand the explanation document or communicate verbally and clearly with the research team. People experiencing homelessness were defined as those who had slept in places other than their homes over the past 30 days. This included individuals sleeping in public spaces such as sidewalks, bus stations, markets, under bridges, and parks, as well as in workplaces, hammock cafés, internet cafés, and guest houses.

### Sampling

For slum dwellers, local collaborators contacted individuals who met the criteria in their neighborhoods to assess their willingness to participate. In each site, a local collaborator was selected from among those who lived in the area, were familiar with the study population, and had a good relationship with them. Those who expressed a willingness to participate were included in a numbered list of potential participants. Simple random sampling was applied using a random number table to recruit participants from this list. For individuals experiencing homelessness, all those contacted by the SCDI team—either through outreach or drop-in services—who agreed to participate in the study were included. In total, 415 individuals with unstable housing were selected for the study, comprising 383 slum dwellers and 32 homeless individuals, and data were collected using a questionnaire.

### Data collection

A structured questionnaire was used, which included the Kessler Psychological Distress Scale (K6) score to identify SPD and questions to elaborate on the following statuses: basic socio-demographic information (age, gender, school years, whether the individual is a migrant or was born in HCMC, marital status, household size, and number of children by age group), basic socio-economic status (means of earning a living, monthly income, ownership of a private house, and sleeping location in the last 30 days), entitlement to social support due to designated vulnerability status, cash or in-kind support received, identification and registration, health insurance coverage, experiences of labor exploitation, incarceration, detention, debt, the impact of and social protection during the COVID-19 pandemic, and HIV and tuberculosis status. The survey was conducted from November 2023 to April 2024.

### Measurement of serious psychological distress

SPD was assessed using the K6 scale, a brief, six-item self-report tool that evaluates general psychological distress experienced over the past 30 days. The scale includes questions about feelings of nervousness, hopelessness, restlessness, depression that cannot be alleviated, excessive effort in daily activities, and feelings of worthlessness. Each item is scored on a 5-point Likert scale, ranging from 0 (none of the time) to 4 (all of the time), yielding a total score between 0 and 24. Higher scores indicate greater distress. A cutoff score of 13 or more was applied, which is commonly used to define SPD [41, 42]. The K6 is widely recognized for its brevity, reliability, and validity in distinguishing cases consistent with DSM-IV diagnoses [43].

### Statistical analysis

We estimated the prevalence of SPD and calculated the proportion of individuals with various socio-economic vulnerabilities, including possession of an ID card, registration at their place of residence, possession of a health insurance card, experiences of any form of labor exploitation, and debt status, using univariate analyses. We then explored the associations between SPD and various explanatory variables presented in the Data Collection section. Chi-square tests were conducted for categorical variables, and t-tests were applied for continuous variables (age, household size, number of children, and monthly income). We constructed a multiple logistic regression model, with SPD as the dependent variable and the explanatory variables that showed significant associations with SPD as independent variables. Statistical significance was set at p < 0.05. Data analyses were performed using SPSS version 27 (IBM Corp., Armonk, NY, USA).

### Ethical considerations

The study protocol, explanation document with the informed consent form, consent withdrawal form, and questionnaire were reviewed and approved by both the NCGM Ethics IRB on July 9, 2023 (NCGM-S-004698-00), and the Ethics IRB of the Institute for Social Development Studies (ISDS), Vietnam, on September 28, 2023, before data collection commenced. Written informed consent with participants’ signatures was obtained from all participants prior to their involvement in the study. The explanation document included the purpose, methods, risks, and benefits of the study. Participants were informed that consent was at their discretion, that they would not be treated unfavorably if they chose not to participate, and that they could withdraw their consent at any time, even after the initial agreement. Copies of the explanation document and consent forms were provided to the participants, while the original consent forms were retained by SCDI. All researchers involved in this study adhered to the Declaration of Helsinki (Revised 2013 Fortaleza) and conducted the research in accordance with the “Ethical Guidelines for Medical and Biological Research Involving Human Subjects” set forth by the Ministry of Health, Labour and Welfare (MOHLW), the Ministry of Education, Culture, Sports, Science and Technology (MEXT), and the Ministry of Economy, Trade and Industry (METI) of Japan.

## Results

### Basic characteristics of study subjects

Table 1 depicts the basic social, demographic, and economic characteristics of the 415 study subjects. The average age was 44.12 years (SD: 0.648). Among the age groups, those aged 50-59 years were the most prevalent, comprising 108 subjects (26.0%), followed by the 40-49 age group with 96 subjects (23.1%). The gender distribution was female-dominant, with females accounting for 244 subjects (58.8%). A total of 383 subjects (92.3%) were slum dwellers, while 32 subjects (7.7%) were homeless. Of the participants, 219 (52.8%) were migrants from other provinces, whereas 196 (47.2%) were born in HCMC. Average duration of schooling was 5.93 years. Additionally, 101 subjects (24.3%) were married, and 79 (19.0%) were cohabiting without formal marriage. The average household size was 4.06 persons (SD: 0.093). The average monthly income was 3,279,164 Vietnamese Dong (VND) (SD: 146,600), which is equivalent to 134.45 US dollars as of February 2024, the midpoint of the survey. Regarding means of earning a living, 104 subjects (25.1%) had no income and were dependent on others; 85 (20.4%) were engaged in informal employment; 58 (14.0%) were garbage collectors; 39 (9.4%) were motorcycle drivers or shippers; 29 (7.0%) were lottery ticket sellers; and 26 (6.3%) were street vendors.

**Table 1.** Basic characteristics of the study subjects.

| Characteristics | Figures |
| --- | --- |
| Total number of study subjects | 415 |
| Age, mean (SD) in years | 44.12 (0.648) |
| Female gender, N (%) | 244 (58.8) |
| Housing status |  |
| Slum dweller, N (%) | 383 (92.3) |
| Homeless, N (%) | 32 (7.7) |
| Migrant or born in HCMC |  |
| Migrant, N (%) | 219 (52.8) |
| Born in HCMC, N (%) | 196 (47.2) |
| Duration of schooling, mean (SD) in years | 5.93 (3.859) |
| Marital status |  |
| Married, N (%) | 101 (24.3) |
| Never married, N (%) | 101 (24.3) |
| Cohabiting without formal marriage, N (%) | 79 (19.0) |
| Separated, divorced or widowed, N (%) | 37 (8.9) |
| Household size, mean (SD) in persons | 3.06 (1.90) |
| Monthly income, mean (SD) in VND | 3,279,164 (2,982,875) |
| Means of earning a living |  |
| No income and dependent on others, N (%) | 104 (25.1) |
| Informal employment | 85 (20.4) |
| Garbage collector | 58 (14.0) |
| Motorcycle driver/shipper | 39 (9.4) |
| Lottery ticket seller | 29 (7.0) |
| Street vender | 26 (6.3) |
| House maid | 13 (3.1) |
| Construction worker | 12 (2.9) |
| Shopkeeper | 12 (2.9) |
| Formal employment | 10 (2.4) |
| Others | 27 (6.5) |

SD: standard deviation, VND: Vietnamese Dong.

### Status of socio-economic vulnerabilities

Table 2 presents the socio-economic vulnerabilities of the study subjects, with a breakdown between slum dwellers and the homeless. A lack of civil registration, evident in the absence of ID cards and registration at their living locations, was particularly prevalent among the homeless population. Health insurance coverage extended to only 58% of the subjects, and just 34% of the homeless. Nearly 70% experienced some form of labor exploitation, and 44% were in debt.

**Table 2.** Status of socio-economic vulnerabilities of the 415 study subjects.

|  | No. of subjects | Prevalence (%) | 95% CI |
| --- | --- | --- | --- |
| **Lack of ID card** |  |  |  |
| All subjects | 49/415 | 11.8 | 8.7-14.9 |
| Slum dwellers | 37/383 | 9.7 | 6.7-12.6 |
| Homeless | 12/32 | 37.5 | 19.8-55.2 |
| **Lack of registration at living location** |  |  |  |
| All subjects | 114/415 | 27.5 | 23.2-31.8 |
| Slum dwellers | 88/383 | 23.0 | 18.7-27.2 |
| Homeless | 26/32 | 81.3 | 67.0-95.5 |
| **Lack of health insurance card** |  |  |  |
| All subjects | 175/415 | 42.2 | 37.4-46.9 |
| Slum dwellers | 154/383 | 40.2 | 35.3-45.1 |
| Homeless | 21/32 | 65.6 | 48.2-83.0 |

| **Faced any form of labor exploitation** |  |  |  |
| --- | --- | --- | --- |
| All subjects | 286/415 | 68.9 | 64.4-73.4 |
| Slum dwellers | 264/383 | 68.9 | 64.3-73.6 |
| Homeless | 22/32 | 68.8 | 51.8-85.7 |
| **In debt** |  |  |  |
| All subjects | 181/415 | 43.6 | 38.8-48.4 |
| Slum dwellers | 168/383 | 43.9 | 38.9-48.9 |
| Homeless | 13/32 | 40.6 | 22.6-58.6 |

CI: confidence interval

Among the 286 subjects who experienced labor exploitation, 211 (73.8%) faced contract rejections, 83 (29.0%) were forced to work excessively hard, 75 (26.2%) experienced wage theft, 53 (18.5%) suffered physical or verbal abuse, and 29 (10.1%) encountered work conditions that did not align with what was promised. Among the 181 subjects who were in debt, 160 (88.4%) borrowed from money lenders with interest, while 21 (11.6%) borrowed from acquaintances without interest. Regarding the reasons for being in debt, 94 (51.9%) borrowed for rent payments, 89 (49.2%) for purchasing food, 72 (39.8%) for covering electricity and water bills, 37 (20.4%) for medical treatments, and 34 (18.8%) for educational expenses for their children.

### Estimated prevalence of SPD

As shown in Table 3, the estimated prevalence of SPD, defined as a K6 score of ≥13, among all 415 subjects was 19.8%. The prevalence among slum dwellers was 18.5%, while among the homeless it was 34.4%.

**Table 3.** Prevalence of serious psychological distress (SPD) among all subjects, slum dwellers, and homeless.

|  | No. of subjects with SPD | Prevalence (%) | 95% CI |
| --- | --- | --- | --- |
| All subjects | 82/415 | 19.8 | 16.0-23.9 |
| Slum dwellers | 71/383 | 18.5 | 14.8-22.8 |
| Homeless | 11/32 | 34.4 | 18.6-53.2 |

CI: confidence interval

### Factors associated with SPD

Table 4 summarizes both bivariate and multiple logistic regression analyses of the association between SPD and various social, demographic, and economic explanatory variables. The bivariate analysis revealed a statistically significant association between SPD and ten independent variables. Among the socio-demographic factors, female gender (p = 0.001), being homeless (p = 0.031), and sleeping in public places (such as on pavements, at bus stations, in markets, or under bridges/parks) (p = 0.000) were significantly associated with SPD. Among the vulnerability factors, individuals living with HIV who can no longer work (p = 0.012), those meeting vulnerability criteria for monthly cash social assistance (p = 0.012), those who have experienced any form of labor exploitation (p = 0.024), and those currently in debt (p = 0.000) were significantly associated with SPD. In terms of civil registration status and public service entitlements, a lack of registration at the current place of residence (p = 0.009), a lack of an ID card (p = 0.009), and a lack of a valid health insurance card (p = 0.019) were significantly associated with SPD. The multiple logistic regression model, which designated SPD as the dependent (outcome) variable and included the ten variables that showed statistically significant associations with SPD in the bivariate analysis (p < 0.05) as independent (explanatory) variables, revealed that only female gender (p = 0.000), experience of any form of labor exploitation (p = 0.046), and being currently in debt (p = 0.000) remained significantly associated with SPD (p < 0.05) after adjusting for confounding.

**Table 4:**
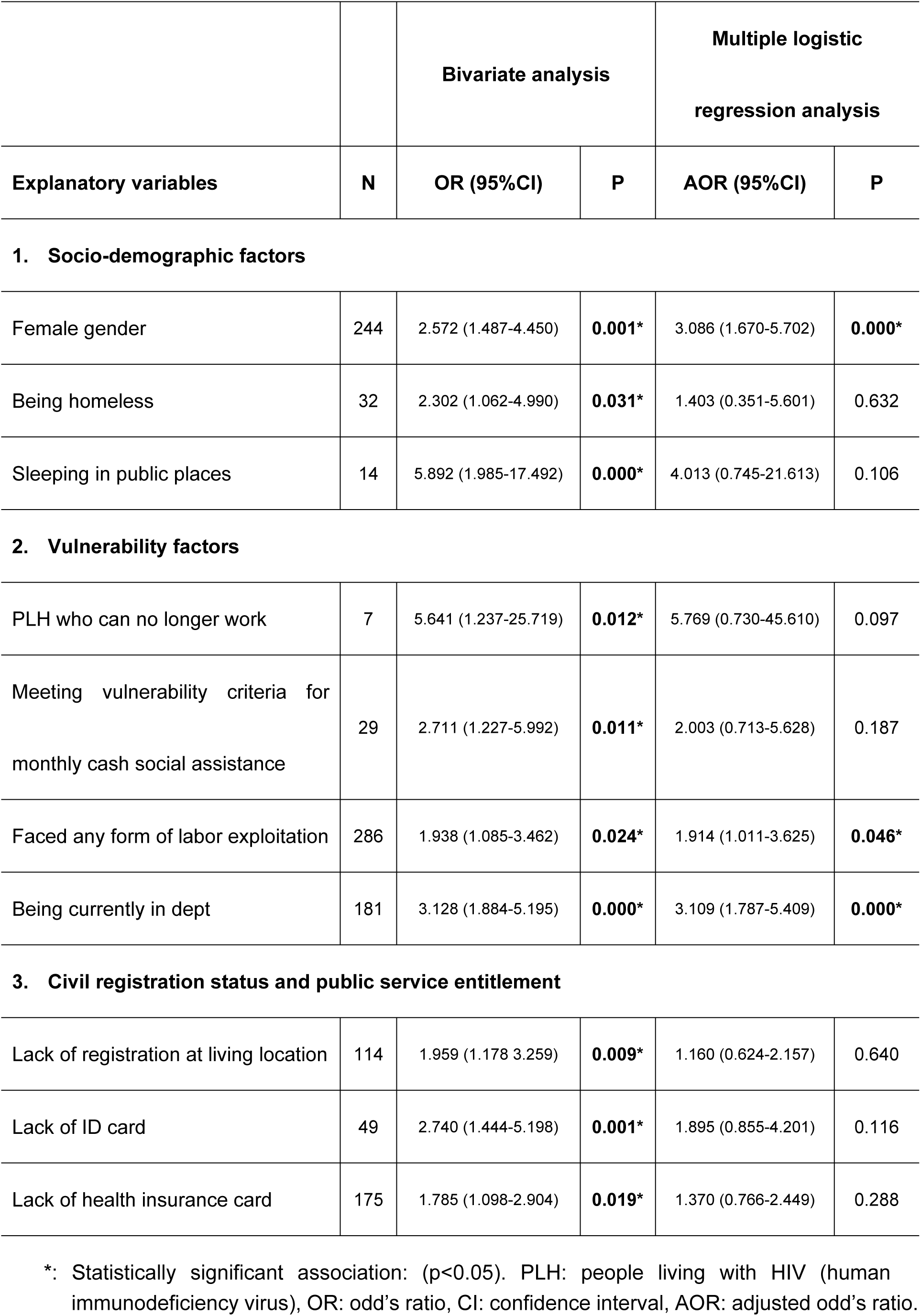
Bivariate and multivariate analyses of associations between serious psychological distress (SPD) and various explanatory variables among the 415 study subjects.

|  |  | **Bivariate analysis** |  | **Multiple logistic regression analysis** |  |
| --- | --- | --- | --- | --- | --- |
| **Explanatory variables** | **N** | **OR (95%CI)** | **P** | **AOR (95%CI)** | **P** |
| 1. Socio-demographic factors |  |  |  |  |  |
| Female gender | 244 | 2.572 (1.487-4.450) | **0.001\*** | 3.086 (1.670-5.702) | **0.000\*** |
| Being homeless | 32 | 2.302 (1.062-4.990) | **0.031\*** | 1.403 (0.351-5.601) | 0.632 |
| Sleeping in public places | 14 | 5.892 (1.985-17.492) | **0.000\*** | 4.013 (0.745-21.613) | 0.106 |
| 2. Vulnerability factors |  |  |  |  |  |
| PLH who can no longer work | 7 | 5.641 (1.237-25.719) | **0.012\*** | 5.769 (0.730-45.610) | 0.097 |
| Meeting vulnerability criteria for monthly cash social assistance | 29 | 2.711 (1.227-5.992) | **0.011\*** | 2.003 (0.713-5.628) | 0.187 |
| Faced any form of labor exploitation | 286 | 1.938 (1.085-3.462) | **0.024\*** | 1.914 (1.011-3.625) | **0.046\*** |
| Being currently in dept | 181 | 3.128 (1.884-5.195) | **0.000\*** | 3.109 (1.787-5.409) | **0.000\*** |
| 3. Civil registration status and public service entitlement |  |  |  |  |  |
| Lack of registration at living location | 114 | 1.959 (1.178-3.259) | **0.009\*** | 1.160 (0.624-2.157) | 0.640 |
| Lack of ID card | 49 | 2.740 (1.444-5.198) | **0.001\*** | 1.895 (0.855-4.201) | 0.116 |
| Lack of health insurance card | 175 | 1.785 (1.098-2.904) | **0.019\*** | 1.370 (0.766-2.449) | 0.288 |

## Discussion

In response to the objective of examining socio-economic vulnerabilities, estimating the prevalence of SPD, and identifying factors associated with SPD among populations with unstable housing, including slum dwellers and homeless individuals in HCMC, Vietnam, the present study revealed precarious civil registration statuses and limited public services entitlement among the subjects. The study estimated the prevalence of SPD at 19.8% (18.5% among slum dwellers and 34.4% among the homeless) and identified female gender, labor exploitation, and indebtedness as factors significantly associated with SPD. These findings make a substantial contribution to both the service delivery and advocacy policies of SCDI and local authorities.

The socio-economic status of our subjects, relative to both national and HCMC standards, was generally disadvantaged. The average monthly income of 3,279,164 VND among our subjects was approximately 66.1% of the national average of 4,962,000 VND and 50.3% of the HCMC average of 6,518,000 VND in 2023 (GSO, Statistical Yearbook 2023). The annualized income of 39 million VND for the subjects exceeded both the national urban poverty line of 24 million VND and the HCMC poverty line of 36 million VND [44]. However, it is noteworthy that nearly a quarter of the subjects had no income. Additionally, because 91.6% of slum dwellers rent rather than own their homes, they must pay rent in addition to utility fees. Often disconnected from kinship and community networks, they lack access to social support, especially when they need to borrow money. As a result, they have to resort to loan sharks when they need funds, and a significant portion of their income goes toward paying interest. The figure for the homeless, at 32 million VND, was below the HCMC poverty line but still above the national urban poverty line. The income poverty rate of our subjects, based on the national urban poverty line, was 37.8%, with almost no difference between the slum dwellers and the homeless, while the nationwide income poverty rate was 4.3% in 2022 [45]. The informal employment rate among our subjects, at 96.8%, far exceeded the national rate of 65.8% and HCMC’s 46.6% in 2023 [16]. Additionally, the unemployment rate among our subjects, at 25.1%, was significantly higher than both the national average of 2.34% and HCMC’s 4.19% in 2023 [16].

The average schooling of our subjects, at 5.93 years, was 3 to 5 years shorter than the national average of 9.4 years in 2022 [16] and the learning-adjusted national average of 10.4 years in 2023 [46]. While 69.2% of household heads nationwide and 62.9% of those in the Southeast Region, including HCMC, were married in 2019 [47], only 43.4% of our subjects— even including those cohabiting without formal marriage—had partners, likely reflecting the precarious living conditions due to unstable housing. The national average household size in 2019 was 3.6 persons [47], while the Southeast Region, including HCMC, averaged 3.3 persons [48]. In contrast, our subjects’ average household size was slightly lower at 3.1 persons, with the figure dropping to 1.4 persons for the homeless.

The precarity of civil registration statuses and limited entitlement to public services among slum dwellers in HCMC has been highlighted by existing literature. A 2012 study of 500 slum households in HCMC described their difficulties in obtaining household registration, land use certificates, and housing certificates due to their temporary residential status. This situation has led to low and unstable incomes, as well as predominantly informal, low-skilled, and precarious employment [17]. Diaz identified three types of slum dwellers in HCMC: 1) long-term urban residents who cannot afford housing outside of slums, 2) rural migrants, and 3) returnees from New Economic Zones (NEZ). Most in categories 1 and 2 are not officially registered to live in HCMC, while NEZ returnees lost their residence permits upon relocation. As a result, the majority of slum dwellers lack legal residential papers, affecting both land tenure and access to public services [49]. A study in two temporary settlements in HCMC revealed a health insurance enrollment rate of 71%, with income being the dominant factor influencing enrollment [44]. Notably, the slums of HCMC are reported to suffer from water, air, and soil pollution, which threaten both the urban ecosystem and the health of the dwellers [19, 50].

The socio-economic vulnerabilities were even more pronounced among the homeless, a finding supported by existing literature. One contributing factor to their unstable lives is the relocation policy. In 2020, the authorities of HCMC began gathering and sending homeless individuals to the HCMC Social Support Center as a measure against the COVID-19 pandemic, with 1,500–2,000 homeless people sent to the center annually [51, 52]. Homeless individuals hiding from authorities were excluded from the list of beneficiaries of the government’s second support package in response to COVID-19 [53]. A qualitative study on homeless youth in HCMC identified barriers they faced in accessing health services, many of which stem from policies that discriminate against the homeless [54]. Several studies have shown that discrimination is associated with PD [35, 55, 56]. When faced with major health issues, homeless youths tended to turn to informal networks—including peers, social workers, and local pharmacies—instead of seeking immediate medical care [54]. Although not included in the scope of this study, street children are also recognized as a major social issue in HCMC [23, 57].

The SPD prevalence of 19.8% among our subjects (18.5% among slum dwellers and 34.4% among the homeless) is significantly higher than those reported in the general population of various countries. In the United States, analyses of National Health Interview Survey (NHIS) data from different periods between 1997 and 2018 revealed that SPD prevalence among the non-institutionalized adult population, as measured by the K6 scale, ranged from 2.7% to 3.4% [58–61]. The 2007 California Health Interview Survey reported an SPD prevalence of 8.6% among adults, also measured by K6 [62]. The Comprehensive Survey of Living Conditions in Japan from 2007 to 2016 found the SPD prevalence among adults, measured by K6, ranged from 4.0% to 4.2% [63]. Various online surveys conducted in Japan during the early phase of the COVID-19 pandemic in 2020 showed SPD prevalence in the general population, as measured by K6, ranging from 9.2% to 11.2% [64–67]. Among undergraduates at Sichuan University in China, the SPD prevalence measured by K6 was 4.0% [68]. The Household, Income and Labor Dynamics in Australia (HILDA) Survey indicated that the SPD prevalence among adults, measured by K10—the 10-item version of the Kessler Psychological Distress Scale—increased from 4.8% in 2007 to 7.4% in 2017 [69]. Published studies on SPD prevalence among the Vietnamese population are very scarce. However, the SPD prevalence among Vietnamese students in the US, as measured by K10, was 18.5%, which is equivalent to the rate found in our slum dweller sample [70].

The prevalence of PD, not limited to SPD, among the Vietnamese general population, as measured by different tools including the K6, SRQ-20, and the Phan Vietnamese Psychiatric Scale (PVPS), ranged from 5.4% to 38.2% [71–74], while figures reported from other countries range from 5% to 27% [75–79]. High PD/SPD prevalence among slum dwellers has been reported. For example, the SPD prevalence among slum dwellers in Port-au-Prince, Haiti, as measured by the K6, was 24.1%, even higher than in our slum dweller sample, while the PD prevalence was 86.5%. Female gender and increasing age were associated with SPD [80]. In São Paulo, Brazil, the PD prevalence among slum dwellers, measured by GHQ-12, was 85.0%, with food insecurity and low income being key contributors to PD [81]. Elevated levels of PD/SPD among the homeless are also highlighted in existing literature. The SPD prevalence among mothers who experienced homelessness and entered shelters between 2010 and 2012 in major US cities, measured by the K6, was 22.0% [82]. In northern California, PD prevalence among homeless adults, measured by CES-D, decreased from 63.7% to 49.4% over a follow-up period of 3 months to 1 year. Securing their own apartment during the follow-up period significantly reduced PD prevalence [83]. Among homeless women in Los Angeles County, the US, the prevalence of mental distress requiring clinical evaluation, as measured by MHI-5, was 45.6% [84]. In the Netherlands, the prevalence of high levels of PD among the homeless in four major cities, as measured by BSI-18, was 39.5%, which decreased to 27.0% after entering a social relief system. Meeting care needs, health insurance coverage, social support from family, and a sense of belonging significantly reduced PD [85].

In line with the findings of the present study, the prevalence of PD is reported to be higher in women than in men in most countries under normal circumstances [35, 79, 86, 87] and during the COVID-19 pandemic [88–90]. Drapeau et al. offer three alternative hypotheses to explain the higher prevalence of PD among women: 1) PD may be partly attributable to gender-related personality traits or biological factors; 2) in most societies, women are either more exposed or more vulnerable to the risk factors associated with PD; and 3) the expression of emotions varies between genders. Nevertheless, despite being the subject of numerous studies, the gender difference in PD remains largely unexplained [35].

Labor exploitation was significantly associated with SPD among the subjects. Abusive (authoritarian and aggressive) supervision has been reported to cause PD in affected workers [91]. Several studies highlight the major impact of violence or harassment at work, either by colleagues or supervisors, on workers’ PD [92–95]. Labor inspections and unionization are potential measures to combat labor exploitation. However, labor inspection in Vietnam faces challenges due to an insufficient number of inspectors, resulting in many enterprises not being adequately inspected [96]. According to the Labor Code, all enterprises are required to establish labor unions [97], but these unions must affiliate with the Vietnam General Confederation of Labor (VGCL), which is linked to the Communist Party [98]. Also, they are often dominated and controlled by employers [99]. Furthermore, two unionist officials from the VGCL and the Ministry of Labor, Invalids, and Social Affairs (MOLISA), who were attempting to align Vietnam’s labor laws with international standards, were arrested in 2024 [100]. These facts raise questions about the feasibility and effectiveness of addressing labor exploitation through unionization. A more feasible approach may lie in the promotion of occupational safety and health (OSH). The Labor Code and Government Decree No. 6 require enterprises to implement all necessary measures for OSH [97]. The Ministry of Health and local health bureaus conduct inspections related to occupational health [97], making this approach an entry point through the health sector, with the potential to influence MOLISA’s chain of command.

Lastly, indebtedness significantly increased the likelihood of SPD, and most of the borrowers were taking loans from moneylenders at interest. Illegal lending practices are common in Vietnam, and loan sharking is often disguised through seemingly lawful moneylenders, pawnshops, social media ads, or street posters. Charging exorbitant interest rates is a crime and is often linked to other illegal activities such as gang involvement, money laundering, or violence [101]. Loan sharks targeting individuals with unstable incomes are prevalent [102]. Law enforcement may be hesitant or unable to investigate or prosecute these moneylenders, as the loans are often made without formal documentation [101]. Despite these challenges, stricter regulation of loan sharks is necessary. In 2020, the Vietnam National Bank issued a circular facilitating debt rescheduling, interest and fee exemptions, or reductions to assist borrowers impacted by the COVID-19 pandemic [52]. Such policies may help reduce the financial and psychological burdens on populations with unstable housing and debt. However, these measures may be difficult to enforce with informal moneylenders. In addition to cracking down on predatory lending, promoting financial inclusion is essential. This is especially important as 26% of our subjects were engaged in small businesses (e.g., selling lottery tickets) and need access to business capital to survive, while approximately 79% of people in Vietnam do not have access to formal financial services [103].

The present study has several limitations. First, since the target population consisted of individuals living in slums or experiencing homelessness, who were accessed by SCDI’s outreach team or sought its drop-in services in HCMC, we cannot generalize the findings to all slum dwellers and the homeless population in HCMC or other areas of Vietnam. Second, we did not comprehensively explore the personal resources aspect of the risk and protective factors related to SPD. To summarize the empirical evidence on the epidemiology of PD, risk and protective factors can be categorized into three groups: 1) sociodemographic factors; 2) stress-related factors; and 3) personal resources [35]. We thoroughly examined categories 1 and 2 in terms of variables related to socio-economic vulnerabilities, including housing status. However, for category 3, we only investigated external resources such as income and education, omitting other external resources like social support and social networks, as well as internal resources such as self-esteem and a sense of control over one’s life.

## Conclusions

The prevalence of SPD among populations with unstable housing, including slum dwellers and the homeless in HCMC, was significantly higher than that of the general population in various countries. SPD was significantly associated with female gender as well as labor and economic exploitation. In terms of labor exploitation, policy interventions, particularly from an occupational health perspective, are necessary. To address economic exploitation through debt, it is essential to implement concurrent efforts to crack down on predatory lending and promote financial inclusion, ensuring access to financial services necessary for maintaining small businesses.

## Data Availability

The data underlying the results presented in the study are available from https://ncgmh-my.sharepoint.com/:u:/g/personal/murakami_it_ncgm_go_jp/EeC1ud4-uh9DnYrpcMfiuPEBFNjIfO82Ohuri6zTSg5qSQ?e=K8BnSy.

https://ncgmh-my.sharepoint.com/:u:/g/personal/murakami_it_ncgm_go_jp/EeC1ud4-uh9DnYrpcMfiuPEBFNjIfO82Ohuri6zTSg5qSQ?e=dhOboC

## Acknowledgements

Authors gratefully acknowledge the contributions of the two community organizations, namely Companions and My Hands, to the planning and implementation of the survey. We also thank Professor Masayoshi Tarui and Mr. Masaki Inaba, who joined in the preliminary survey in HCMC, and their encouragement and inspiration provided to us.

## Notes

### Competing Interest Statement

The authors have declared no competing interest.

### Funding Statement

Yes

### Author Declarations

The study protocol, explanation document with the informed consent form, consent withdrawal form, and questionnaire were reviewed and approved by both the National Center for Global Health and Medicine (NCGM), Japan Ethics IRB on July 9, 2023 (NCGM-S-004698-00), and the Ethics IRB of the Institute for Social Development Studies (ISDS), Vietnam, on September 28, 2023, before data collection commenced.

## References

1 . D’Alessandro D, Appolloni L. Housing and health: an overview. Ann Ig. 2020 October 01;32(5 Supple 1):17–26. doi:10.7416/ai.2020.3391

2 . Mwoka M, Biermann O, Ettman CK, Abdalla SM, Ambuko J, Pearson M, et al. Housing as a social determinant of health: Evidence from Singapore, the UK, and Kenya: The 3-D Commission. Journal of urban health. 2021;98:15–30. doi:10.1007/s11524-021-00557-8

3 . Lancet T. Housing: an overlooked social determinant of health. Lancet (London, England). 2024;403(10438):1723. doi:10.1016/S0140-6736(24)00914-0

4. Taylor L. Housing and health: an overview of the literature. Health Affairs Health Policy Brief. Washington DC: Health Affairs; 2018. Available from: https://www.healthaffairs.org/content/briefs/housing-and-health-overview-literature

5 . Ziersch A, Due C. A mixed methods systematic review of studies examining the relationship between housing and health for people from refugee and asylum seeking backgrounds. Soc Sci Med. 2018;213:199–219. doi:10.1016/j.socscimed.2018.07.045

6 . Downing J. The health effects of the foreclosure crisis and unaffordable housing: a systematic review and explanation of evidence. Soc Sci Med. 2016;162:88–96. doi:10.1016/j.socscimed.2016.06.014

7 . Tsai AC. Home foreclosure, health, and mental health: a systematic review of individual, aggregate, and contextual associations. PloS one. 2015;10(4):e0123182. doi:10.1371/journal.pone.0123182

8 . Vásquez-Vera H, Palència L, Magna I, Mena C, Neira J, Borrell C. The threat of home eviction and its effects on health through the equity lens: a systematic review. Soc Sci Med. 2017;175:199–208. doi:10.1016/j.socscimed.2017.01.010

9 . Jones AA, Gicas KM, Mostafavi S, Woodward ML, Leonova O, Vila-Rodriguez F, et al. Dynamic networks of psychotic symptoms in adults living in precarious housing or homelessness. Psychol Med. 2022;52(13):2559–69. doi:10.1017/s0033291720004444

10 . Myhrvold T, Småstuen MC. The mental healthcare needs of undocumented migrants: an exploratory analysis of psychological distress and living conditions among undocumented migrants in Norway. J Clin Nurs. 2017;26(5-6):825–39. doi:10.1111/jocn.13670

11 . Navarro-Lashayas MA, Eiroa-Orosa FJ. Substance use and psychological distress is related with accommodation status among homeless immigrants. Am J Orthopsychiatry. 2017;87(1):23. doi:10.1037/ort0000213

12 . Patterson ML, Moniruzzaman A, Frankish CJ, Somers JM. Missed opportunities: childhood learning disabilities as early indicators of risk among homeless adults with mental illness in Vancouver, British Columbia. BMJ open. 2012;2(6):e001586. doi:10.1136/bmjopen-2012-001586

13 . Rani D, Singh JK, Acharya D, Paudel R, Lee K, Singh SP. Household food insecurity and mental health among teenage girls living in urban slums in Varanasi, India: a cross-sectional study. International journal of environmental research and public health. 2018;15(8):1585. doi:10.3390/ijerph15081585

14 . Marbin D, Gutwinski S, Schreiter S, Heinz A. Perspectives in poverty and mental health. Frontiers in Public Health. 2022;10:975482. doi:10.3389/fpubh.2022.975482

15 . Carrere J, Vásquez-Vera H, Pérez-Luna A, Novoa AM, Borrell C. Housing insecurity and mental health: the effect of housing tenure and the coexistence of life insecurities. Journal of Urban Health. 2022;99(2):268–76. doi:10.1007/s11524-022-00619-5

16. General Statistics Office, Vietnam. Vietnam Statistical Yearbook 2023. Hanoi: General Statistics Office, Vietnam; 2024. Available from: https://www.gso.gov.vn/wp-content/uploads/2024/06/NIEN-GIAM-THONG-KE-2023_Ban-quyen.pdf

17 . Xoan NTH. Housing statuses of the poor in Ho Chi Minh City, Vietnam. Sociology. 2015;3(2):23–34.

18 . Ooi GL, Phua KH. Urbanization and slum formation. Journal of Urban Health. 2007;84:27–34. doi:10.1007/s11524-007-9167-5

19. Wust S, Bolay J, Du TTN. Metropolization and the ecological crisis: precarious settlements in Ho Chi Minh City, Vietnam. In: Westendorff D, editor. From unsustainable to inclusive cities. Geneva: United Nations Research Institute for Social Development (UNRISD); 2002. pp. 64–79.

20 . Drakakis-Smith D, Dixon C. Sustainable urbanization in Vietnam. Geoforum. 1997;28(1):21–38. doi:10.1016/s0016-7185(97)85525-x

21 . Dang NA. The mega-urban transformations of Ho Chi Minh City in the era of Doi Moi renovation. In: Jones GW, Douglass M, editors. Mega-Urban Regions in Pacific-Asia: Urban Dynamics in a Global Era. Singapore: Singapore University Press; 2008. pp.185–213.

22. Dapice D, Gomez-Ibanez JA, Nguyen XT. Ho Chi Minh City: the challenges of growth. Hanoi: United Nations Development Programme in Vietnam; 2009. Available from: https://www.undp.org/sites/g/files/zskgke326/files/migration/vn/24699_20911_HCM_Challenges_of_growth.pdf

23. Hong D, Ohno K. Street children in Vietnam: interactions of old and new causes in a growing economy (Discussion Paper, No. 6). Hanoi: Vietnam Development Forum; 2005. Available from: https://www.streetchildren.org/wp-content/uploads/2013/03/street-children-in-vietnam.pdf

24. Survey finds only 39 homeless families in HCM City. Viet Nam News Oct 17, 2019 [cited Sep 29, 2024]. Available from: https://vietnamnews.vn/opinion/537035/survey-finds-only-39-homeless-families-in-hcm-city.html

25 . Abdi F, Rahnemaei FA, Shojaei P, Afsahi F, Mahmoodi Z. Social determinants of mental health of women living in slum: a systematic review. Obstetrics & Gynecology Science. 2021;64(2):143–55. doi:10.5468/ogs.20264

26 . Alaazi DA, Menon D, Stafinski T. Health, quality of life, and wellbeing of older slum dwellers in sub-Saharan Africa: a scoping review. Global Public Health. 2021;16(12):1870–88. doi:10.1080/17441692.2020.1840610

27 . Ezeh A, Oyebode O, Satterthwaite D, Chen Y, Ndugwa R, Sartori J, et al. The history, geography, and sociology of slums and the health problems of people who live in slums. The lancet. 2017;389(10068):547–58. doi:10.1016/s0140-6736(16)31650-6

28 . Nejad FN, Ghamari MR, Kamal SHM, Tabatabaee SS, Ganjali R. The most important social determinants of slum dwellers’ health: a scoping review. Journal of Preventive Medicine and Public Health. 2021;54(4):265. doi:10.3961/jpmph.21.073

29 . Hossain MM, Sultana A, Tasnim S, Fan Q, Ma P, McKyer ELJ, et al. Prevalence of mental disorders among people who are homeless: An umbrella review. Int J Soc Psychiatry. 2020;66(6):528–41. doi:10.1177/0020764020924689

30 . Fazel S, Khosla V, Doll H, Geddes J. The prevalence of mental disorders among the homeless in western countries: systematic review and meta-regression analysis. PLoS medicine. 2008;5(12):e225. doi:10.1371/journal.pmed.0050225

31 . Gutwinski S, Schreiter S, Deutscher K, Fazel S. The prevalence of mental disorders among homeless people in high-income countries: an updated systematic review and meta-regression analysis. PLoS medicine. 2021;18(8):e1003750. doi:10.1371/journal.pmed.1003750

32 . Hodgson KJ, Shelton KH, van den Bree MB, Los FJ. Psychopathology in young people experiencing homelessness: A systematic review. Am J Public Health. 2013;103(6):e24–37. doi:10.2105/ajph.2013.301318

33 . Schreiter S, Bermpohl F, Krausz M, Leucht S, Rössler W, Schouler-Ocak M, et al. The prevalence of mental illness in homeless people in Germany: a systematic review and meta-analysis. Deutsches Aerzteblatt International. 2017;114(40):665. doi:10.3238/arztebl.2017.0665

34 . Bassuk EL, Richard MK, Tsertsvadze A. The prevalence of mental illness in homeless children: A systematic review and meta-analysis. Journal of the American Academy of Child & Adolescent Psychiatry. 2015;54(2):86,96. e2. doi:10.1016/j.jaac.2014.11.008

35. Drapeau A, Marchand A, Beaulieu-Prévost D. Epidemiology of psychological distress. In: L’Abate editor. Mental illnesses-understanding, prediction and control. Rijeka, Croatia: InTech; 2011. pp. 105–134. Available from: https://www.intechopen.com/chapters/25512

36 . Ridner SH. Psychological distress: Concept analysis. J Adv Nurs. 2004;45(5):536–45.

37 . Dohrenwend BP, Dohrenwend BS. Perspectives on the past and future of psychiatric epidemiology. The 1981 Rema Lapouse Lecture. Am J Public Health. 1982;72(11):1271–9. doi:10.2105/ajph.72.11.1271

38 . Payton AR. Mental health, mental illness, and psychological distress: Same continuum or distinct phenomena? J Health Soc Behav. 2009;50(2):213–27. doi:10.1177/002214650905000207

39 . Sunderland M, Slade T, Stewart G, Andrews G. Estimating the prevalence of DSM-IV mental illness in the Australian general population using the Kessler Psychological Distress Scale. Australian & New Zealand Journal of Psychiatry. 2011;45(10):880–9. doi:10.3109/00048674.2011.606785

40 . Sakurai K, Nishi A, Kondo K, Yanagida K, Kawakami N. Screening performance of K6/K10 and other screening instruments for mood and anxiety disorders in Japan. Psychiatry Clin Neurosci. 2011;65(5):434–41. doi:10.1111/j.1440-1819.2011.02236.x

41 . Kessler RC, Barker PR, Colpe LJ, Epstein JF, Gfroerer JC, Hiripi E, et al. Screening for serious mental illness in the general population. Arch Gen Psychiatry. 2003;60(2):184–9. doi:10.1001/archpsyc.60.2.184

42 . Kessler RC, Galea S, Gruber MJ, Sampson NA, Ursano RJ, Wessely S. Trends in mental illness and suicidality after Hurricane Katrina. Mol Psychiatry. 2008;13(4):374–84. doi:10.1038/sj.mp.4002119

43 . Kessler RC, Andrews G, Colpe LJ, Hiripi E, Mroczek DK, Normand S, et al. Short screening scales to monitor population prevalences and trends in non-specific psychological distress. Psychol Med. 2002;32(6):959–76. doi:10.1017/s0033291702006074

44. Chi VLT. Urban poverty during COVID-19 in Vietnam: A case study of the Ma Lang-Dong Tien Neighborhood, Ho Chi Minh City, Vietnam. Tokyo: JICA Ogata Research Institute; 2023. Available from: https://www.jica.go.jp/english/jica_ri/publication/discussion/__icsFiles/afieldfile/2023/08/01/Discussion_Paper_No.13.pdf

45. World Bank. Poverty & equity brief, Viet Nam, April 2024. Washington DC: World Bank; 2024. Available from: https://www.worldbank.org/en/topic/poverty/publication/poverty-and-equity-briefs

46. Human Capital Project, World Bank. Human capital country brief, Vietnam (October 2023). Washington DC: World Bank; 2023. Available from: https://thedocs.worldbank.org/en/doc/64e578cbeaa522631f08f0cafba8960e-0140062023/related/HCI-AM23-VNM.pdf

47. General Statistics Office, Vietnam. Completed results of the 2019 Viet Nam population and housing census. Hanoi: General Statistics Office, Vietnam; 2020. Available from: https://www.gso.gov.vn/wp-content/uploads/2019/12/Ket-qua-toan-bo-Tong-dieu-tra-dan-so-va-nha-o-2019.pdf

48. Central Population and Housing Census Steering Committee, Vietnam. The Viet Nam population and housing census of 00:00 hours on 1 April 2019: Implementation organisation and preliminary results. Hanoi: Central Population and Housing Census Steering Committee, Vietnam; 2020. Available from: https://www.gso.gov.vn/wp-content/uploads/2019/10/2.-ENG_Census-on-Housing-and-Population_2019_final.pdf

49. Diaz CA. Temporary upgrading: How permanent are the results? A case study of strategies to improve tenure in Ho Chi Minh City. MCP. Thesis, Massachusetts Institute of Technology. 2002. Available from: https://dspace.mit.edu/bitstream/handle/1721.1/65730/50854787-MIT.pdf?sequence=2&isAllowed=y

50 . Bolay J, Cartoux S, Cunha A, Du TTN, Bassand M. Sustainable development and urban growth: precarious habitat and water management in Ho Chi Minh City, Vietnam. Habitat International. 1997;21(2):185–97. doi:10.1016/s0197-3975(97)89956-0

51. HCM City to keep beggars, homeless out of harm’s way amid pandemic. Viet Nam News. Apr 3, 2020 [cited Sep 29, 2024]. Available from: https://vietnamnews.vn/society/674597/hcm-city-to-keep-beggars-homeless-out-of-harm-s-way-amid-pandemic.html

52. Habitat for Humanity Vietnam. Input for report on COVID-19 and right to housing. Hanoi: Habitat for Humanity Vietnam;2020. Available from: https://www.ohchr.org/sites/default/files/Documents/Issues/Housing/COVID19/CivilSociety/HabitatVietnam.docx

53. United Nations Development Programme in Vietnam. Rapid assessment of the COVID-19: Socio-economic impact on vulnerable households in Vietnam. Hanoi: United Nations Development Programme in Vietnam; 2021. Available from: https://www.undp.org/vietnam/publications/rapid-assessment-covid-19-socio-economic-impact-vulnerable-households-viet-nam

54 . Boggiano VL, Harris LM, Schmidt V, Nguyen HA, Barry M. Protecting, balancing, and confronting: Health seeking among homeless youth in Ho Chi Minh City, Vietnam. International Journal of Child, Youth and Family Studies. 2017;8(3/4):1–25. doi:10.18357/ijcyfs83/4201717998

55 . González-Castro JL, Ubillos S. Determinants of psychological distress among migrants from Ecuador and Romania in a Spanish city. Int J Soc Psychiatry. 2011;57(1):30–44. doi:10.1177/0020764009347336

56 . Yip T, Gee GC, Takeuchi DT. Racial discrimination and psychological distress: the impact of ethnic identity and age among immigrant and United States-born Asian adults. Dev Psychol. 2008;44(3):787. doi:10.1037/0012-1649.44.3.787

57. Terre des Hommes. A study on street children in Ho Chi Minh City. Ho Chi Minh City: Terre des Hommes; 2001.

58. Weissman JS, Pratt LA, Miller EA, Parker JD. Serious psychological distress among adults, United States, 2009-2013 (NCHS Data Brief No. 203). Hyattsville, MD: National Center for Health Statistics (NCHS), Centers for Disease Control and Prevention (CDC); 2015. Available from: https://www.cdc.gov/nchs/data/databriefs/db203.pdf

59 . Pratt LA, Dey AN, Cohen AJ. Characteristics of adults with serious psychological distress as measured by the K6 scale, United States, 2001-04. Advance data from vital and health statistics no. 382. Hyattsville, MD: National Center for Health Statistics (NCHS), Centers for Disease Control and Prevention (CDC);.2007. Available from: https://citeseerx.ist.psu.edu/document?repid=rep1&type=pdf&doi=0a79504a88e039e2a67113d7877b64ef534f348e

60 . Pratt LA. Serious psychological distress, as measured by the K6, and mortality. Ann Epidemiol. 2009;19(3):202–9. doi:10.1016/j.annepidem.2008.12.005

61. Daly M. Prevalence of psychological distress among working-age adults in the United States, 1999–2018. Am J Public Health. 2022;112(7):1045–9. doi:10.2105/ajph.2022.306828

62 . Prochaska JJ, Sung H, Max W, Shi Y, Ong M. Validity study of the K6 scale as a measure of moderate mental distress based on mental health treatment need and utilization. International journal of methods in psychiatric research. 2012;21(2):88–97. doi:10.1002/mpr.1349

63 . Nishi D, Susukida R, Usuda K, Mojtabai R, Yamanouchi Y. Trends in the prevalence of psychological distress and the use of mental health services from 2007 to 2016 in Japan. J Affect Disord. 2018;239:208–13. doi:10.1016/j.jad.2018.07.016

64 . Kikuchi H, Machida M, Nakamura I, Saito R, Odagiri Y, Kojima T, et al. Development of severe psychological distress among low-income individuals during the COVID-19 pandemic: longitudinal study. BJPsych Open. 2021;7(2):e50. doi:10.1192/bjo.2021.5

65 . Kikuchi H, Machida M, Nakamura I, Saito R, Odagiri Y, Kojima T, et al. Changes in psychological distress during the COVID-19 pandemic in Japan: a longitudinal study. Journal of Epidemiology. 2020;30(11):522–8. doi:10.2188/jea.je20200271

66 . Yoshioka T, Okubo R, Tabuchi T, Odani S, Shinozaki T, Tsugawa Y. Factors associated with serious psychological distress during the COVID-19 pandemic in Japan: a nationwide cross-sectional internet-based study. BMJ open. 2021;11(7):e051115. doi:10.1136/bmjopen-2021-051115

67 . Okubo R, Yoshioka T, Nakaya T, Hanibuchi T, Okano H, Ikezawa S, et al. Urbanization level and neighborhood deprivation, not COVID-19 case numbers by residence area, are associated with severe psychological distress and new-onset suicidal ideation during the COVID-19 pandemic. J Affect Disord. 2021;287:89–95. doi:10.1016/j.jad.2021.03.028

68 . Kang Y, Guo W, Xu H, Chen Y, Li X, Tan Z, et al. The 6-item Kessler psychological distress scale to survey serious mental illness among Chinese undergraduates: Psychometric properties and prevalence estimate. Compr Psychiatry. 2015;63:105–12. doi:10.1016/j.comppsych.2015.08.011

69 . Butterworth P, Watson N, Wooden M. Trends in the prevalence of psychological distress over time: comparing results from longitudinal and repeated cross-sectional surveys. Frontiers in Psychiatry. 2020;11:595696. doi:10.3389/fpsyt.2020.595696

70 . Quynh NN, Linh LM. Mental distress and coping strategies of Vietnamese students in US during the COVID-19 pandemic. TNU Journal of Science and Technology. 2022; 228(03):13–20. doi:10.34238/tnu-jst.6525

71 . Hoang MT, Do KN, Pham HQ, Nguyen CT, Ha GH, Vu GT, et al. Psychological distress among mountainous farmers in Vietnam: a cross-sectional study of prevalence and associated factors. BMJ open. 2020;10(8):e038490. doi:10.1136/bmjopen-2020-038490

72 . Bao Giang K, Viet Dzung T, Kullgren G, Allebeck P. Prevalence of mental distress and use of health services in a rural district in Vietnam. Global Health Action. 2010;3(1):2025. doi:10.3402/gha.v3i0.2025

73 . Richardson LK, Amstadter AB, Kilpatrick DG, Gaboury MT, Tran TL, Trung LT, et al. Estimating mental distress in Vietnam: the use of the SRQ-20. Int J Soc Psychiatry. 2010;56(2):133–42. doi:10.1177/0020764008099554

74 . Rees S, Silove D, Chey T, Steel Z, Bauman A, Phan T. Physical activity and psychological distress amongst Vietnamese living in the Mekong Delta. Australian & New Zealand Journal of Psychiatry. 2012;46(10):966–71. doi:10.1177/0004867412459568

75 . Benzeval M, Judge K. Income and health: the time dimension. Soc Sci Med. 2001;52(9):1371–90. doi:10.1016/s0277-9536(00)00244-6

76 . Chittleborough CR, Winefield H, Gill TK, Koster C, Taylor AW. Age differences in associations between psychological distress and chronic conditions. International journal of public health. 2011;56:71–80. doi:10.1007/s00038-010-0197-5

77 . Gispert R, Rajmil L, Schiaffino A, Herdman M. Sociodemographic and health-related correlates of psychiatric distress in a general population. Soc Psychiatry Psychiatr Epidemiol. 2003;38:677–83. doi:10.1007/s00127-003-0692-6

78 . Kuriyama S, Nakaya N, Ohmori-Matsuda K, Shimazu T, Kikuchi N, Kakizaki M, et al. Factors associated with psychological distress in a community-dwelling Japanese population: the Ohsaki Cohort 2006 Study. Journal of epidemiology. 2009;19(6):294–302. doi:10.2188/jea.je20080076

79 . Phongsavan P, Chey T, Bauman A, Brooks R, Silove D. Social capital, socio-economic status and psychological distress among Australian adults. Soc Sci Med. 2006;63(10):2546–61. doi:10.1016/j.socscimed.2006.06.021

80 . Tymejczyk O, Rivera VR, Mireille P, Dorelien A, Petion JS, Grace S, et al. Psychological distress among a population-representative sample of residents of four slum neighborhoods in Port-au-Prince, Haiti. J Affect Disord. 2020;263:241–5. doi:10.1016/j.jad.2019.11.103

81 . Santana C, Manfrinato CV, Souza P, Marino A, Condé VF, Stedefeldt E, et al. Psychological distress, low-income, and socio-economic vulnerability in the COVID-19 pandemic. Public Health. 2021;199:42–5. doi:10.1016/j.puhe.2021.08.016

82. Shinn M, Gubits D, Dunton L. Behavioral health improvements over time among adults in families experiencing homelessness (Homeless families research brief, OPRE report no. 2018-61). Rockville, MD: Abt Associates. 2018. Available from: https://www.acf.hhs.gov/sites/default/files/documents/opre/opre_behavioral_health_brief_09_06_2018_508.pdf

83 . Wong YI. Tracking change in psychological distress among homeless adults: An examination of the effect of housing status. Health Soc Work. 2002;27(4):262–73. doi:10.1093/hsw/27.4.262

84 . Austin EL, Andersen R, Gelberg L. Ethnic differences in the correlates of mental distress among homeless women. Womens Health Issues. 2008;18(1):26–34. doi:10.1016/j.whi.2007.08.005

85 . Van Straaten B, Rodenburg G, Van der Laan J, Boersma SN, Wolf JR, Van de Mheen D. Changes in social exclusion indicators and psychological distress among homeless people over a 2.5-year period. Soc Indicators Res. 2018;135:291–311. doi:10.1007/s11205-016-1486-z

86 . Caron J, Liu A. Factors associated with psychological distress in the Canadian population: a comparison of low-income and non low-income sub-groups. Community Ment Health J. 2011;47:318–30. doi:10.1007/s10597-010-9306-4

87 . Jorm AF, Windsor TD, Dear K, Anstey KJ, Christensen H, Rodgers B. Age group differences in psychological distress: the role of psychosocial risk factors that vary with age. Psychol Med. 2005;35(9):1253–63. doi:10.1017/s0033291705004976

88 . Xiong J, Lipsitz O, Nasri F, Lui LM, Gill H, Phan L, et al. Impact of COVID-19 pandemic on mental health in the general population: A systematic review. J Affect Disord. 2020;277:55–64. doi:10.1016/j.jad.2020.08.001

89 . Alkhamees AA, Alrashed SA, Alzunaydi AA, Almohimeed AS, Aljohani MS. The psychological impact of COVID-19 pandemic on the general population of Saudi Arabia. Compr Psychiatry. 2020;102:152192. doi:10.1016/j.comppsych.2020.152192

90 . Cao W, Fang Z, Hou G, Han M, Xu X, Dong J, et al. The psychological impact of the COVID-19 epidemic on college students in China. Psychiatry Res. 2020;287:112934. doi:10.1016/j.psychres.2020.112934

91 . Tepper BJ. Consequences of abusive supervision. Academy of management journal. 2000;43(2):178–90. doi:10.5465/1556375

92 . Marchand A, Demers A, Durand P. Does work really cause distress? The contribution of occupational structure and work organization to the experience of psychological distress. Soc Sci Med. 2005;61(1):1–14. doi:10.1016/j.socscimed.2004.11.037

93 . Mueller CW, De Coster S, Estes SB. Sexual harassment in the workplace: Unanticipated consequences of modern social control in organizations. Work Occup. 2001;28(4):411–46. doi:10.1177/0730888401028004003

94 . Piotrkowski CS. Gender harassment, job satisfaction, and distress among employed white and minority women. J Occup Health Psychol. 1998;3(1):33. doi:10.1037//1076-8998.3.1.33

95 . Richman JA, Rospenda KM, Nawyn SJ, Flaherty JA, Fendrich M, Drum ML, et al. Sexual harassment and generalized workplace abuse among university employees: prevalence and mental health correlates. Am J Public Health. 1999;89(3):358–63. doi:10.2105/ajph.89.3.358

96. Pham TTH. The development of capacity for labour inspection: a case study of Ministry of Labour, Invalids and Social Affairs in Vietnam. M.Sc. Thesis, University of Tampere. 2018. Available from: https://core.ac.uk/download/pdf/250152482.pdf

97. Qi L, Taylor B, Frost S. Labour relations and regulation in Vietnam: Theory and practice. Hong Kong: Southeast Asia Research Centre, City University of Hong Kong; 2003. Available from: https://www.cityu.edu.hk/searc/Resources/Paper/WP53_03_QiTaylorFrost.pdf

98. Fair Labor Association. Independent external monitoring report of SanMar, Vietnam. Washington DC: Fair Labor Association; 2008. Available from: https://core.ac.uk/download/pdf/33605888.pdf

99 . Trinh L. Trade union organizing free from employers’ interference: Evidence from Vietnam. Southeast Asian Studies. 2014;3(3):589–609. doi:10.20495/seas.3.3_589

100. Strangio S. Vietnam arrests second labor activist in a month, rights group claims. The Diplomat. May 22, 2024 [cited Sep 29, 2024]. Available from: https://thediplomat.com/2024/05/vietnam-arrests-second-labor-activist-in-a-month-rights-group-claims/

101. Department of Foreign Affairs and Trade, Australia. DFAT Country Information Report Vietnam. Canberra: Department of Foreign Affairs and Trade, Australia; 2022. Available from: https://www.dfat.gov.au/sites/default/files/country-information-report-vietnam.pdf

102. Vietnam central bank enhances measures against loan sharks. VIetnamnet Global. Jun 5, 2019 [cited Sep 29, 2024]. Available from: https://vietnamnet.vn/en/vietnam-central-bank-enhances-measures-against-loan-sharks-539055.html

103. Independent Advisory Group on Country Information (IAGCI). Country policy and information note Vietnam: Fear of illegal moneylenders. London: Independent Advisory Group on Country Information (IAGCI); 2023. Available from: https://assets.publishing.service.gov.uk/media/63c6b784d3bf7f580416e78e/VNM_CPIN_Fear_of_illegal_money_lenders.pdf

